# Identifying type 1 and 2 diabetes in population level data: assessing the accuracy of published approaches

**DOI:** 10.1101/2022.04.11.22273617

**Authors:** Nicholas J Thomas, Andrew McGovern, Katherine G Young, Seth A Sharp, Michael N Weedon, Andrew T Hattersley, John Dennis, Angus G Jones

## Abstract

**Aims:** Population datasets are increasingly used to study type 1 or 2 diabetes, and inform clinical practice. However, correctly classifying diabetes type, when insulin treated, in population datasets is challenging. Many different approaches have been proposed, ranging from simple age or BMI cut offs, to complex algorithms, and the optimal approach is unclear. We aimed to compare the performance of approaches for classifying insulin treated diabetes for research studies, evaluated against two independent biological definitions of diabetes type.

**Method:** We compared accuracy of thirteen reported approaches for classifying insulin treated diabetes into type 1 and type 2 diabetes in two population cohorts with diabetes: UK Biobank (UKBB) n=26,399 and DARE n=1,296. Overall accuracy and predictive values for classifying type 1 and 2 diabetes were assessed using: 1) a type 1 diabetes genetic risk score and genetic stratification method (UKBB); 2) C-peptide measured at >3 years diabetes duration (DARE).

**Results:** Accuracy of approaches ranged from 71%-88% in UKBB and 68%-88% in DARE. All approaches were improved by combining with requirement for early insulin treatment (<1 year from diagnosis). When classifying all participants, combining early insulin requirement with a type 1 diabetes probability model incorporating continuous clinical features (diagnosis age and BMI only) consistently achieved high accuracy, (UKBB 87%, DARE 85%). Self-reported diabetes type alone had high accuracy (UKBB 87%, DARE 88%) but was available in just 15% of UKBB participants. For identifying type 1 diabetes with minimal misclassification, using models with high thresholds or young age at diagnosis (<20 years) had the highest performance. An online tool developed from all UKBB findings allows the optimum approach of those tested to be selected based on variable availability and the research aim.

**Conclusion:** Self-reported diagnosis and models combining continuous features with early insulin requirement are the most accurate methods of classifying insulin treated diabetes in research datasets without measured classification biomarkers.

## Background

### Robustly classifying diabetes type in population level data is challenging

Large population level datasets are widely used for clinical studies of people with diabetes, however for results to be robust, accurate diabetes classification is fundamental. Together type 1 diabetes (T1D) and type 2 diabetes (T2D) account for ≥98% of all diabetes cases, [1] but these two subtypes have marked differences in aetiology, pathophysiology and management [2]. While absence of insulin treatment in longstanding diabetes is highly specific for T2D [2, 3], classifying currently insulin treated diabetes cases is challenging [3]. The clinical diagnosis of insulin treated diabetes cases is frequently not available in population data, and if available will include substantial misclassification and miscoding (≈15%)[4-8]. In population data, biomarkers which can help improve classification, such as C-peptide or islet autoantibodies [9], are rarely available.

### The comparative performance of approaches to classify insulin treated diabetes in epidemiological studies is unknown

The optimum approach for classifying T1D and T2D in population data remains unclear. Previously published approaches vary and include: clinician or self-reported diabetes type, diabetes treatment, billing codes or using specific cut offs of diabetes related features for example body mass index (BMI) or age at diabetes diagnosis [10-21]. Where the performance of these approaches has been assessed, it is normally against a clinical assessment of T1D or T2D diagnosis [7, 10, 11, 16-21]. These assessments will not only suffer from the inaccuracies of clinical diagnosis and coding, but also a circularity bias that features favored by clinicians for determining diabetes type will appear to be most discriminatory. While prediction models for classification have been developed and tested against C-peptide and histology defined diabetes type these have not been compared to other approaches [12, 15, 22]. To date, there has not been an evaluation of the comparative performance of existing classification approaches against a robust independent biomarker.

### Aim

To help researchers to choose the optimum diabetes classification approach for research studies using large datasets, we aimed to compare the performance of a number of published approaches for classifying insulin treated diabetes in two population level datasets. Classification approaches were evaluated against two independent biological definitions of diabetes type based on type 1 diabetes genetic risk scores (T1GRS), and measured C-peptide.

## Method

Within two population cohorts we assessed the performance of different published approaches for classifying insulin treated diabetes into T1D and T2D against biomarker defined diabetes subtypes. In UK Biobank we used a type 1 diabetes genetic risk score (T1DGRS) within a previously published genetic stratification method [23] to compare the proportion of T1D and T2D cases correctly and incorrectly classified by each approach. We also assessed the performance of these approaches in a large unselected population cohort with diabetes (the DARE cohort) against diabetes type defined by C-peptide level measured after a median 14 years duration.

### Study design and participants

#### UK Biobank

UK Biobank recruited a population cohort of more than 500,000 people aged between 40 and 70 years registered with the UK National Health Service [24]. We evaluated a subset of 26,399 unrelated individuals self-reporting diabetes. To allow direct comparison of classification approaches in the same cohort, individuals were excluded where missing BMI measurement (n=237) or self-reported age at diabetes diagnosis (n=1,675). A further 1,389 participants were excluded where it was not possible to generate a T1DGRS. Overall 23,098 participants met study eligibility criteria, a study flowchart is shown in electronic supplementary materials (ESM) Figure 1a. A subset of 45% (10,491/23,098) of participants had linkage to their primary care record.

The main analysis was restricted to the 72% (16,619/23,098) of participants of white European descent, as the T1DGRS used to define diabetes type has not been validated in non-white ethnicities [25, 26]. People of white European descent were those who self-identified as white European and were confirmed as ancestrally white by use of principal components analyses of genome-wide genetic information [27]. A separate secondary exploratory analysis was undertaken including all 23,098 participants of all ethnicities. Clinical history was self-reported via an interactive questionnaire and nurse led interview, further details of clinical features and lipid assessment are given in ESM.

### DARE cohort

The DARE study recruited, predominantly though primary care in the South West of England, an unselected population of adults with diabetes (regardless of age of onset or diabetes type, gestational diabetes excluded) [5]. We evaluated 1,296 participants (22% (1296/5991) of the DARE cohort) with measured C-peptide. C-peptide was measured on stored non fasting EDTA at DARE recruitment after January 2010 as previously described (see ESM) [5]. Participants were excluded where BMI measurement was missing (n=6) or if diabetes duration at recruitment was ≤3 years (n=49) due to the limitations of C-peptide assessment in short duration diabetes [9]. A study flow chart is shown in ESM Figure 1b. Although all ethnicities were recruited to DARE, 99% were white (1224/1241). In DARE all clinical history was self-reported by participants in an interview with a research nurse as reported previously [5].

### Assessment of population approaches for classifying diabetes type in insulin treated individual

Overall we compared ten different approaches for the classification of insulin treated diabetes with the variables required for each approach listed in Table 1. For all approaches using continuous variables, cut offs to classify either T1D or T2D were selected based on previously proposed values where available [7, 10-12, 15]. Different cut offs were used where the aim was to classify all insulin treated participants or select a T1D or T2D cohort with minimal misclassification (Table 1). The approaches evaluated were: age at diagnosis alone ((approach 1: *age*), BMI alone (approach 2: *BMI*), a T1D probability clinical features model, incorporating BMI and age at diagnosis (approach 3: *clinical model*) [12, 22], a T1D probability model incorporating BMI and age at diagnosis, sex, HDL, triglycerides and total cholesterol (approach 4: *lipid model*) [15], a tree structured model using hospital episode statistics (HES) data including ICD10 and ICD9 diabetes codes and history of diabetic ketoacidosis (DKA) (approach 5: *ICD Algorithm*) [18] and an algorithm developed for classifying diabetes type within UK Biobank (approach 6: *Biobank algorithm*) [10]. The UK Biobank algorithm defines diabetes as probable or possible T1D or T2D using three different flow charts incorporating age at diagnosis, time to Insulin, non-metformin oral hypoglycaemic agents (OHA), self-report of type 1 diabetes and ethnicity. In our study the UK Biobank algorithm was evaluated with and without the addition of self-reported diabetes type. For the model incorporating lipids, in the 26% (1262/4845) of UK Biobank participants where lipids were not available, instead of excluding these cases, their calculated clinical model probability as above (BMI and age at diagnosis) was substituted, allowing classification of all participants. While both the clinical and lipid models can also incorporate islet antibodies and T1DGRS this was not assessed due to lack of islet autoantibodies and the use of T1DGRS for outcome definition in UK biobank, and the performance incorporating these markers in DARE being already published [12, 15]. Self-reported diabetes type as a single marker was also evaluated (approach 7: *Self*).

**Table 1:**
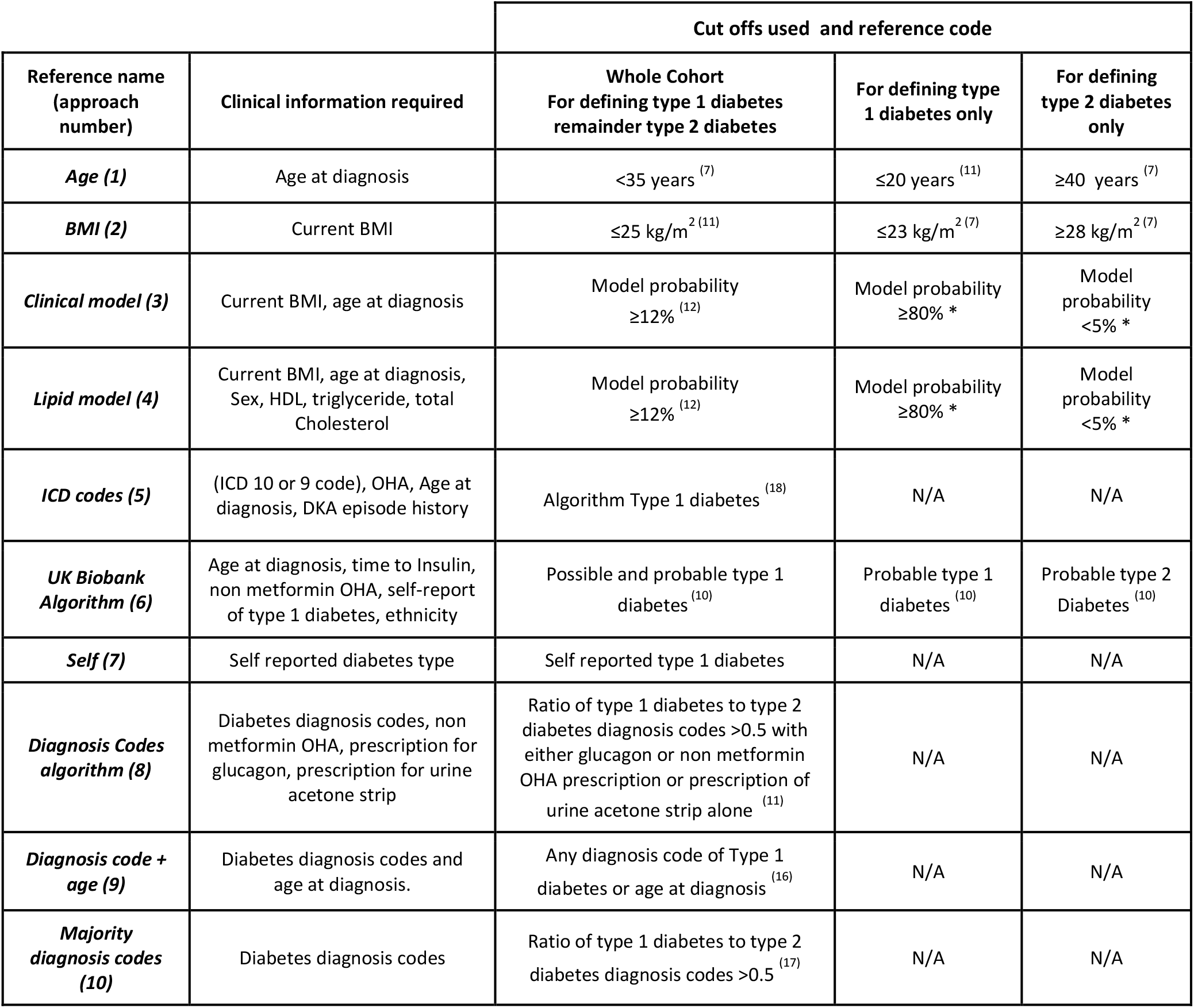
Diabetes specific factors required for each approach and the different cut offs required for classifying all cases, or defining type 1 diabetes or type 2 diabetes. Where available cut offs were taken from existing literature [7, 10-12, 16-18]. * For the models previously published cut offs were not available for selecting pure T1D and T2D cohorts so pragmatic values were chosen from published data aiming for 100% and >90% PPV for T1D classification and 100% PPV for T2D classification [12].

In the subset of UK Biobank participants with linked primary care data, recorded diagnostic codes were used to classify diabetes type, within the following approaches: a classification algorithm incorporating diagnosis codes and prescriptions for non-metformin OHA, glucagon or urine acetone (ketone) strips [11] ((approach 8: *diagnosis codes algorithm*), diagnosis codes used alone [17] (approach 9: *diagnostic codes alone*) and diagnostic code in combination with age of diagnosis [16] (approach 10: *diagnosis codes and age*). Linked primary care data were not available in DARE. The code for all approaches is provided in ESM appendix 2.

For identifying ‘pure’ type 1 and 2 diabetes using prediction models, no previous cut off has been recommended, therefore cut offs were chosen prior to analysis based on probability thresholds that gave high positive predictive value (PPV) for type 1 or 2 diabetes in previous literature [12]: T1D ≥80% probability and T2D <5% probability, for defining T1D a further cut-off of 20% probability was evaluated to give a high PPV whilst aiming to capture a high percentage of all T1D cases. Rapid insulin requirement and OHA treatment are well reported to associate with T1D and T2D respectively. Therefore as an additional analysis performance of approaches was further evaluated with the addition of knowledge of rapid insulin requirement defined as insulin treatment within a year of diagnosis, or also by current treatment with any OHA.

### Biological definitions of diabetes type approaches evaluated against

#### UK Biobank

We have recently shown that measuring the average polygenic susceptibility to T1D (captured by a T1D genetic risk score (T1DGRS)) of a population with diabetes can allow the proportion of T1D in that population to be determined [23, 28]. Genetic risk cannot be used in isolation to define diabetes type at an individual level as a high genetic susceptibility for T1D does not prevent a person having T2D [29]. However, within a cohort the average distribution of T1DGRS will reflect the proportion of T1D to T2D cases within it. A T1D cohort defined by an accurate classification approach will have a high average genetic susceptibility to T1D, closely mirroring the genetic risk of a reference cohort with confirmed T1D [3]. Conversely a T1D cohort defined by an inaccurate classification approach will have a lower average genetic susceptibility to T1D, as it will also include T2D cases with neutral genetic susceptibility for T1D. This concept allows the proportion of T1D to T2D within cohorts defined by different classification approaches to be estimated and the relative performance of different approaches to be compared. The main analysis was restricted to participants of white European descent, where the T1DGRS is validated, with a separate secondary exploratory analysis undertaken in all ethnicities.

We generated a 67 SNP type 1 diabetes T1DGRS as previously described [30], see ESM methods for further detail.

### DARE

T1D was defined as severe insulin deficiency: measured non-fasting C-peptide <200pmol/L. Type 2 diabetes was defined as participants currently insulin treated with a C-peptide ≥200pmol/L. All analysed participants had a duration of diabetes at C-peptide measurement of over three years [9]. C-peptide was measured on stored non-fasted EDTA at DARE recruitment and as part of routine care ESM and as previously described [5].

### Statistical analysis

When classifying all insulin treated cases, approaches were ranked by the overall accuracy of each definition, defined as the proportion of all T1D and T2D cases correctly classified relative to the total number of all cases classified. For each approach the PPV of cases called T1D and T2D (percent of those identified who have the condition as defined by the biological standard) and sensitivity for detecting T1D and T2D (percentage of cases with the condition identified) were also calculated. Where aiming to classify just a T1D or T2D cohort, approaches were ranked firstly based on PPV and then secondly by sensitivity.

### UK Biobank

For each classification approach the mean T1DGRS for cases classified as T1D (*Approach*_*Called T*1*D*_) and T2D (*Approach*_*Called T*2*D*_) were separately evaluated against mean T1DGRS for reference T1D cases (*Reference*_*T*1*D*_) (n=6483 mean T1DGRS = 14.50) and reference Type 2 diabetes equivalent cohort (*Reference*_*T*2*D*_) (n= 9246 mean T1DGRS = 10.37) both taken from the Type 1 Diabetes Genetics Consortium [31]. Reference T1D cases were white European, clinically diagnosed and aged <17 years at diagnosis. The higher the proportion of diabetes cases correctly defined by a classification approach the more the T1DGRS of the groups classified as T1D or T2D will respectively genetically resemble true T1D and T2D reference populations (method shown in ESM figure 2). The proportion of T1D within groups, defined by each classification approach, is then calculated according to the normalised difference of each clinical definitions mean T1DGRS (*Approach*_*Called* (*T*1*D*/*T*2*D*)_) and the mean T1DGRS of the two reference populations (*Reference*_*T*1*D*_ and *Reference*_*T*2*D*_) in the equations below and as described previously [28, 32]. For cases defined as having T1D by each classification approach, PPV for T1D is equivalent to *Proportion*_*T*1*D*_. For cases defined as having T2D by each classification approach, positive predictive value (PPV) for T2D is calculated as 1-*Proportion* _*T*1*D*_.

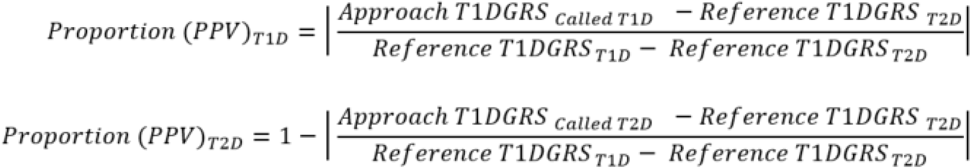

### Calculating accuracy in UK Biobank and DARE

Where all insulin treated participants were classified as having either T1D or T2D, accuracy was calculated as:

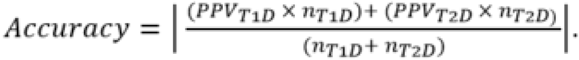

Where *n*_*T*1*D*_ is the number of cases called as having T1D and *n*_*T*2*D*_ is the number of cases called as having T2D by each approach.

All analyses were performed using Stata 16 (StataCorp LP, College Station, TX).

## Results

### Performance of approaches to classifying all insulin treated white European participants with diabetes in UK Biobank

Within the UK Biobank, of the white European participants meeting eligibility criteria, 21% (3534/16,619) were insulin treated. The clinical characteristics of all participants split by insulin treatment status are shown in ESM Table 1. In the 13,085 participants with diabetes not currently insulin treated the mean T1DGRS (10.32, SD 2.38) was consistent with a classical non-T1D reference population [31] mean T1DGRS (10.37 SD 2.26) suggesting little to no T1D in this group. The genetically assessed performance of classification approaches to classify all insulin treated diabetes cases as either T1D or T2D ranked by accuracy are shown in Table 2.

**Table 2:**
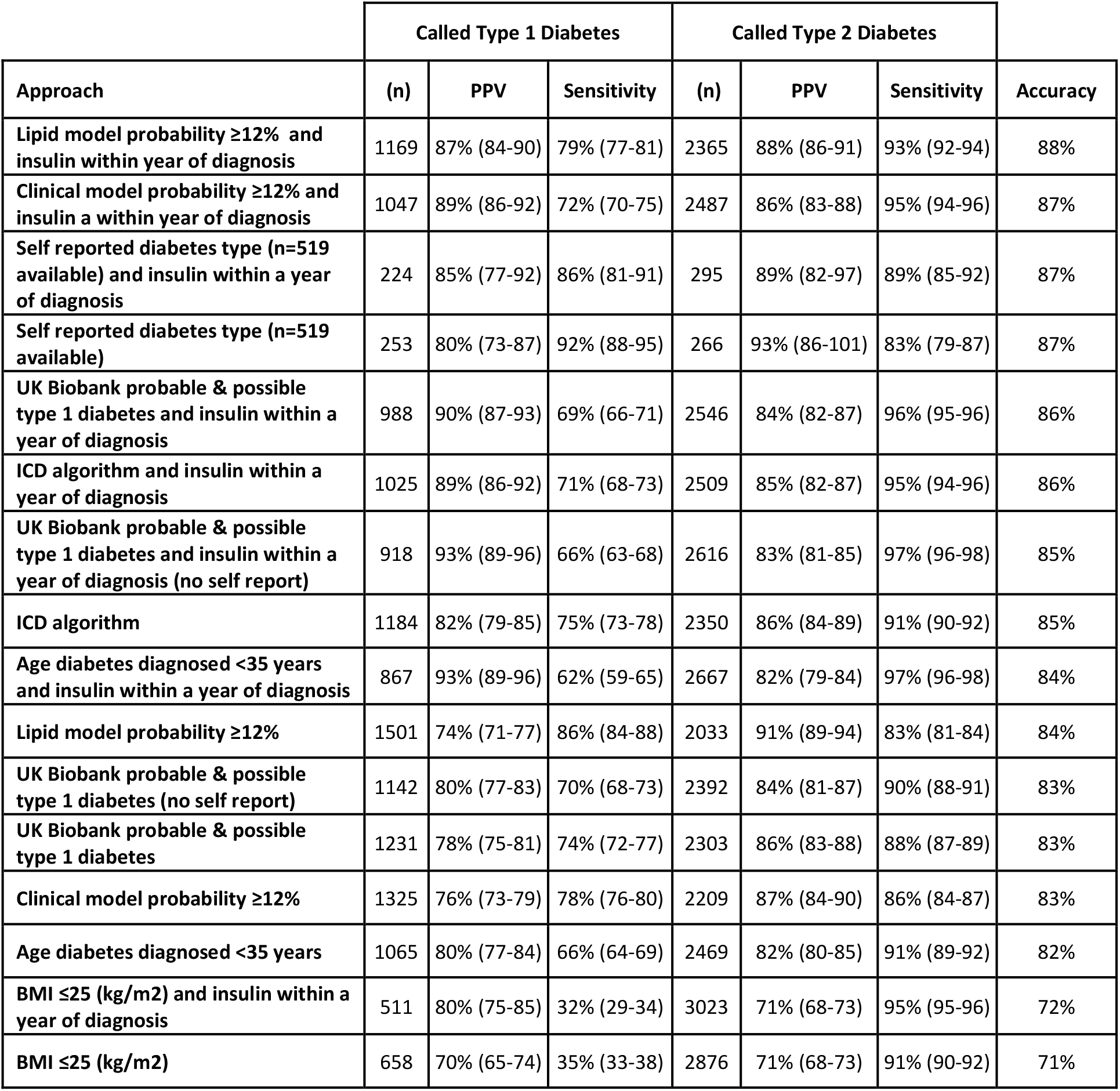
Comparative performance of approaches classifying all insulin treated white European participants with diabetes in UK Biobank. *Cases are classified as type 1 diabetes if they meet the stated criteria, and are otherwise classified as type 2 diabetes. Results ranked by accuracy (total correctly classified) then type 1 diabetes PPV. Brackets signify 95% CI*, Positive predictive value (PPV).

The median classification accuracy was 85%, and varied substantially by approach (range 71% to 88%). The highest accuracy overall was rapid insulin requirement combined with the *clinical model*, overall correctly classifying 87% and *lipid model* overall correctly classifying 88%. Self-reported diabetes type with or without the addition of rapid insulin had accuracy of 87% but was available in just 15% (519/3534) of all cases. For the majority of approaches, adding rapid insulin treatment requirement to define T1D substantially improved accuracy with absence of OHA treatment only slightly less accurate, ESM table 2. The lowest accuracy was seen in approaches using simple cut-offs for individual variables: *age of diagnosis (<35 years) 82%* and *BMI (≤25kg/m2) 71%*. In the 47% (1644/3534) of the insulin treated cohort with linked primary care data, diabetes *diagnosis codes algorithm* alongside rapid insulin requirement gave the highest accuracy of approaches that incorporate electronic healthcare record data and diagnosis codes at 85%. For direct comparison ESM table 3 gives the performance of other classification approaches in this reduced subset of the dataset with linked primary care records, results were broadly similar in this subset, again models with and without lipids, combined with early insulin treatment having the highest overall accuracy of 88% and 87% respectively.

**Table 3:** Comparative performance of approaches classifying all insulin treated participants with diabetes in DARE. *Cases are classified as type 1 diabetes if they meet the stated criteria, and are otherwise classified as type 2 diabetes. Results ranked by accuracy (total correctly classified) then type 1 diabetes PPV. Brackets signify 95% CI*, Positive predictive value (PPV). Lipid model and Diagnosis codes not evaluated as unavailable in DARE.

|  | Called Type 1 Diabetes |  |  | Called Type 2 Diabetes |  |  |  |
| --- | --- | --- | --- | --- | --- | --- | --- |
| Approach | (n) | PPV | Sensitivity | (n) | PPV | Sensitivity | Accuracy |
| Self reported diabetes type and insulin within a year of diagnosis | 310 | 89% (85-92) | 82% (78-86) | 474 | 88% (85-91) | 92% (90-95) | 88% |
| Self reported diabetes type | 335 | 86% (83-90) | 87% (83-90) | 449 | 90% (87-93) | 90% (87-93) | 88% |
| UK Biobank probable & possible type 1 diabetes and insulin within a year of diagnosis | 325 | 86% (82-90) | 84% (80-88) | 459 | 88% (85-91) | 90% (87-93) | 87% |
| Clinical model probability $\geq 12\%$ and insulin a within year of diagnosis | 278 | 90% (86-93) | 75% (70-79) | 506 | 83% (80-86) | 94% (91-96) | 85% |
| UK Biobank probable & possible type 1 diabetes and insulin within a year of diagnosis (no self report) | 257 | 90% (87-94) | 69% (65-74) | 527 | 81% (77-84) | 94% (92-97) | 84% |
| UK Biobank probable & possible type 1 diabetes | 392 | 76% (72-80) | 89% (86-92) | 392 | 91% (88-93) | 79% (75-83) | 83% |
| Age diabetes diagnosed <35 years and insulin within a year of diagnosis | 242 | 90% (86-94) | 65% (60-70) | 542 | 79% (75-82) | 95% (93-97) | 82% |
| Clinical model probability $\geq 12\%$ | 346 | 78% (74-82) | 81% (77-85) | 438 | 85% (82-89) | 83% (80-87) | 82% |
| UK Biobank probable & possible type 1 diabetes (no self report) | 300 | 81% (77-85) | 73% (68-78) | 484 | 81% (78-85) | 87% (84-90) | 81% |
| Age diabetes diagnosed <35 years | 280 | 80% (76-85) | 67% (62-72) | 504 | 78% (75-82) | 88% (85-91) | 79% |
| BMI $\leq 25$ (kg/m<sup>2</sup>) and insulin within a year of diagnosis | 140 | 87% (82-93) | 37% (31-42) | 644 | 67% (63-71) | 96% (94-98) | 71% |
| BMI $\leq 25$ (kg/m<sup>2</sup>) | 187 | 72% (66-79) | 40% (35-46) | 597 | 67% (63-70) | 88% (85-91) | 68% |

### Performance of approaches to classifying all insulin treated participants with diabetes in DARE

In the DARE cohort we identified 1241 people with diabetes who met our inclusion criteria, 63% (784/1241) were insulin treated with 42% (333/784) having a C-peptide <200 pmol/l consistent with T1D, at a median duration of 18 years. Table 3 gives the performance of classification approaches to classify all insulin treated diabetes cases as either T1D or T2D against a C-peptide definition of diabetes type. Accuracy values and overall ranking of approaches were similar to when diabetes type was defined genetically in UK Biobank, median accuracy 83% (range 68%-88%). In DARE the clinical model combined with rapid insulin requirement had accuracy of 85%. Self-reported diabetes type alone gave the highest accuracy 88%. The Biobank algorithm (incorporating self-reported diabetes type) with rapid insulin had accuracy of 87%. This reduced to 84% when self-reported diabetes type was not included within the algorithm. Again all methods were improved by adding rapid insulin requirement. In the 451 non-insulin treated participants with C-peptide measured 99.6% (449/451) had a C-peptide ≥200 consistent with T2D.

### Performance of approaches to optimally identify type 1 diabetes amongst insulin treated participants with diabetes

The performance of methods to optimally identify T1D, ranked by PPV in UK Biobank (percent of those identified as T1D who have the condition genetically) are shown in table 4. A pure T1D cohort was generated when rapid insulin requirement was combined with either age at diagnosis ≤ 20 years (PPV 100%) or a clinical model probability ≥80% (PPV 99%). However, these approaches had low sensitivity respectively only identifying 33% and 37% of all T1D cases. Using probable T1D in the Biobank algorithm combined with rapid insulin requirement identified 69% of all T1D cases, with a PPV of 90%. This was similar to using a lower clinical model probability of ≥20% identifying 67% of all T1D cases with a PPV of 91%. Comparable results for the majority of approaches for both PPV and sensitivity of T1D identified were achieved in DARE, using C-peptide defined diabetes type, Table 4.

**Table 4:** Comparative performance of approaches classifying type 1 diabetes in UK Biobank and DARE in insulin treated participants. Cases are classified as type 1 diabetes if they meet the stated criteria, and are otherwise classified as type 2 diabetes. Results ranked in UK Biobank by type 1 diabetes PPV then sensitivity for identifying type 1 diabetes. Analysis in UK Biobank restricted to White Europeans. Brackets signify 95% CI, Positive predictive value (PPV).

|  | UK BIOBANK |  | DARE |  |
| --- | --- | --- | --- | --- |
| Approach | PPV of cases called type 1 diabetes | Sensitivity for identifying type 1 diabetes | PPV of cases called type 1 diabetes | Sensitivity for identifying type 1 diabetes |
| Age diabetes diagnosed $\leq 20$ years and insulin within a year of diagnosis | 100% (99-100) | 33% (30-35) | 96% (93-100) | 40% (32-49) |
| Clinical model probability $\geq 80\%$ and insulin within a year of diagnosis | 99% (98-100) | 37% (34-39) | 96% (93-99) | 47% (39-54) |
| Lipid model probability $\geq 80\%$ and insulin within a year of diagnosis | 97% (95-98) | 40% (38-43) | n/a | n/a |
| Lipid model probability $\geq 80\%$ | 92% (90-94) | 42% (39-45) | n/a | n/a |
| Age diabetes diagnosed $\leq 20$ years | 92% (90-95) | 34% (31-36) | 96% (92-99) | 40% (32-49) |
| Clinical model probability $\geq 20\%$ and insulin within a year of diagnosis | 91% (89-92) | 67% (65-70) | 91% (88-95) | 70% (65-76) |
| Clinical model probability $\geq 80\%$ | 91% (88-93) | 37% (35-40) | 93% (90-97) | 47% (39-55) |
| UK Biobank probable type 1 diabetes and insulin within a year of diagnosis | 90% (88-92) | 69% (66-71) | 86% (82-90) | 84% (80-88) |
| Lipid model probability $\geq 20\%$ and insulin within a year of diagnosis | 89% (87-91) | 75% (73-77) | n/a | n/a |
| UK Biobank probable type 1 diabetes | 89% (87-91) | 70% (67-72) | 84% (80-88) | 88% (84-91) |
| BMI $\leq 23$ (kg/m<sup>2</sup>) and insulin within a year of diagnosis | 82% (78-87) | 16% (14-18) | 90% (83-97) | 19% (10-28) |
| Self reported type 1 diabetes | 80% (75-85) | 92% (88-95) | 86% (83-90) | 87% (83-90) |
| Lipid model probability $\geq 20\%$ | 80% (78-82) | 81% (79-84) | n/a | n/a |
| Clinical model probability $\geq 20\%$ | 80% (78-83) | 71% (69-74) | 84% (80-88) | 74% (69-79) |
| BMI $\leq 23$ (kg/m<sup>2</sup>) | 75% (70-80) | 17% (15-19) | 79% (70-87) | 20% (11-28) |

#### Performance of approaches to optimally identify type 2 diabetes within insulin treated participants with diabetes

Performance of methods to optimally identify T2D, ranked by PPV in UK Biobank (percent of those identified as T2D who have the condition genetically) are shown in ESM table 4. A pure T2D cohort was generated using probable T2D within the Biobank Algorithm, PPV 100% but this had low sensitivity capturing just 17% of all insulin treated T2D cases. A clinical model probability <5% gave a T2D PPV of 94% and captured 67% of all T2D cases. Adding absence of rapid insulin requirement to all definitions of T2D increased T2D PPV in all approaches but resulted in a lower proportion of all T2D cases being captured. Comparable results for both PPV and sensitivity for each approach were achieved in DARE, ESM Table 4.

#### Performance of approaches to classifying all insulin treated participants with diabetes in UK Biobank

As an exploratory analysis we evaluated the performance of approaches to classify all participants with diabetes in UK Biobank regardless of ethnicity. Within the 4,845 insulin treated participants the overall performance of approaches was similar to when undertaken in just White Europeans, median accuracy 85% (range 75% to 91%) with the best accuracy achieved using probability models combined with rapid insulin requirement: lipid model 91%, clinical features only model 90%, ESM Table 5.

#### Development of algorithm for optimal approach selection

We developed a pragmatic online tool for researchers to select the optimum approach of those evaluated for classifying insulin treated diabetes cases in population datasets, based on findings in UK Biobank: <u>Classifying Diabetes for Research: Method Selector (newcastlerse.github.io)</u>. The optimal approach varies based on the research question being asked and the diabetes outcomes available in the dataset being used. ESM appendix 2 provides researchers the R code to implement all methods which is also provided within the online tool.

## Discussion

We evaluated the performance of approaches for classifying diabetes type in two different population level datasets: UK Biobank and DARE. Results were consistent across datasets despite using two different biological definitions of diabetes type. The impact of classification approach selection on study results and conclusions is highlighted by the marked variation in accuracy observed in our study. Across the two different datasets combining rapid insulin requirement with T1D models incorporating BMI and age at diagnosis (*clinical model*) and these features with lipids (*lipid model)* consistently achieved the highest accuracy for classifying all insulin treated participants (≥85%). Self-reported diabetes type showed similar accuracy in both UK Biobank and the DARE cohort but was only recorded in the minority (15%) of UK Biobank participants limiting its utility.

Our results suggest that when available self-reported diabetes type can be used to classify population cohorts with insulin treated diabetes. When self-reported diabetes type is unavailable, as seen in UK Biobank in the majority of cases, probability models combined with rapid insulin requirement provide a highly accurate simple alternative. Approaches combining different variables had higher accuracy than cut-offs applied to single variables such as BMI or age at diagnosis. All approaches are improved by adding variables capturing either rapid insulin requirement or current OHA treatment. It was possible to identify pure T1D cohorts in both datasets though use of a combination of early insulin treatment and either high model probability or very young age at diagnosis.

A key strength of our study was that performance was evaluated against biological definitions of diabetes type. This reduces the potential for inaccuracies and bias if testing against clinical definitions which are subject to both error and circularity (with features accurate for clinical classification reflecting features clinicians consider to be important) [4, 5, 7]. The main analysis in UK Biobank was restricted to white European participants where the T1DGRS has been validated. As an exploratory analysis we evaluated all participants to show that the ranking of approaches remained similar (meaning the optimum approach remains valid) even if the absolute accuracy of approaches in all non-white European ethnicities should be interpreted with caution. Whilst all ethnicities were included in DARE 99% of participants were white European.

Few studies have compared different classification methods to robust biomarker defined diabetes types. In a cohort with insulin treated diabetes, Hope et al evaluated the performance of age of diagnosis <35 to classify diabetes cases with T1D defined by C-peptide deficiency and cases with preserved C-peptide defined as T2D [7]. Age at diagnosis correctly classified 83% of all cases in their study comparable with in our study: 82% in UK Biobank and 79% in DARE. This remained comparable when age of diagnosis was combined with rapid insulin requirement: Hope et al study accuracy of 85% versus 84% UK Biobank and 82% DARE. Model performance was also high when previously assessed against diabetes subtypes defined by pancreatic histology in the NPOD consortium [22]. Because knowledge of time to insulin treatment is not always available, we also evaluated absence of oral hypoglycemic treatment to define T1D showing this was only slightly less accurate than rapid insulin requirement. The importance of insulin treatment in helping initially determine diabetes type in population datasets is emphasized by the genetic susceptibility of all participants not currently insulin treated being consistent with little to no T1D in this group. In DARE absence of insulin treatment was also almost never associated with C-peptide deficiency despite use of a pragmatic random sample, mirroring previous studies defining diabetes type using C-peptide [33].

Limitations of our study include that T1DGRS is known to modestly reduce with increasing age of T1D diagnosis, [34-37] and our T1D reference T1DGRS cohort were all diagnosed with T1D under 17 years of age [30]. In previous studies the mean T1DGRS of those with confirmed type 1 diabetes diagnosed over 30 years of age was 2.5% lower than those diagnosed <18 years [38]. In UK Biobank the majority (90% (3194/3534)) of insulin treated participants were aged >17 years at diagnosis, making our genetic estimates of T1D proportions slight underestimates. However, as all approaches were evaluated within the same dataset the comparative performance results remain robust and reassuringly similar results were also found defining diabetes type by C-peptide in DARE. Both the Biobank algorithm (developed in UK Biobank) and the T1D clinical model (developed in a cohort that included DARE) were evaluated in the same cohorts they were developed in. Reassuringly both methods performed comparatively well in the alternative data set they were not developed in suggesting any bias was minimal. Despite using both T1D probability models in all participants even though they were developed in adults aged 18-50 they were consistently high performing approaches in both datasets [12, 15]. It is possible accuracy could have been further improved by varying cutoffs in older adults however this would have risked being over fitted. Lipids in UK Biobank were also unfasted, in contrast to the model development dataset, and it is therefore possible performance would increase where fasted lipids are available [15]. Using genetic predisposition to T1D can be helpful in diabetes classification; in the original development of the clinical model adding T1DGRS improved performance [12] and we would recommend using this when genetic data is available, however as T1DGRS was our outcome we were unable to evaluate this approach. Islet autoantibodies used in combination with clinical models also improve performance [12], but are rarely available in population data as is the case in UK Biobank. Classifying diabetes as only being T1D or T2D will miss other types of diabetes. Reassuringly in DARE just 2% (29/1241) of the cohort had a clinician diagnosis which was not T1D or T2D. Finally we have only tested one of a number of published algorithms for using primary care electronic healthcare records (available only in a subset of UK Biobank participants, and unavailable in the DARE cohort). The approach proposed by Klompas using primary care data [11] has been extensively used, and therefore was included as an example of an approach that makes use of the additional data available in electronic healthcare records.

Our results are important for all researchers studying type 1 or 2 diabetes. The considerable differences in pathophysiology, treatment and associated risks of T1D and T2D means inadvertently studying mixed cohorts could lead to misleading study findings [39]. Our results allow determination of the optimal approach for classifying insulin treated diabetes cases whilst also confirming that non-insulin treated cases of over three years duration can confidently be labelled as having T2D. Approaches can be selected based on which diabetes specific outcomes are available and the research question being asked. An added advantage of our study is that researchers can understand the accuracy of the approach used and how this might impact their results and their relatability to other studies where different approaches may have been used. For ease our findings have been translated into an online tool allowing researchers to determine and then implement the optimal approach for their research question and dataset.

## Conclusion

Within two separate population datasets and using two different biological definitions of diabetes we show the performance of approaches for classifying insulin treated diabetes type for research studies and translate this into an online tool for optimal approach selection for researchers. Self-reported diagnosis and models combining continuous features are the most accurate methods of classifying insulin treated diabetes in research datasets without measured classification biomarkers.

## Data Availability

UK Biobank data are available through a procedure described at http://www.ukBiobank.ac.uk/using-the-resource/.
DARE data are available through application to the Peninsula Research Bank https://exetercrfnihr.org/about/exeter-10000-prb/

## Contributors

NJT, AM and AGJ designed the study. SAS, KGY and MNW acquired the data and SAS and MNW generated the T1DGRS. NJT, JD, AM and AGJ analysed the data. NJT wrote the first draft of the report. All authors reviewed the draft, contributed to the revision of the report and gave final approval for publication. AGJ and NJT are the guarantors of this work.

## Acknowledgments

The authors thank participants who took part in the study and the research teams who undertook cohort recruitment.

This research has in part been conducted using UK Biobank Resource. Biobank application 9055.

The authors are grateful to Mike Simpson of Newcastle University for developing the online tool.

## Data Availability

UK Biobank data are available through a procedure described at http://www.ukBiobank.ac.uk/using-the-resource/.

DARE data are available through application to the Peninsula Research Bank https://exetercrfnihr.org/about/exeter-10000-prb/

