## Supplementary material for "Identifying type 1 and 2 diabetes in population level data: assessing the accuracy of published approaches": supplmentary

**Electronic Supplementary material (ESM)**

**ESM methods**

In UK Biobank the following diabetes features were self-reported via an interactive questionnaire completed at baseline: age of diagnosis, insulin use within one year of diagnosis and insulin use at enrolment to the study. Diabetes type, current treatment with insulin and treatment with oral hypoglycemic agents (OHA) were recorded during a research nurse led interview. Participants were recorded as being currently insulin treated if insulin treatment was either self-reported or recorded at interview. In DARE all clinical history was self-reported by participants in an interview with a research nurse.

Body mass index (BMI) in UK Biobank and DARE was calculated as (weight [kg]/height $[m]^2$) recorded at the recruitment visit.

**Lipids**

In UK Biobank baseline Total Cholesterol, HDL and Triglycerides measures were performed on non-fasted serum blood samples. Sample handling and analysis has previously been described (1; 2). No adjustment was made for lipid treatment status. Lipids were unavailable in the majority (79% (1028/1296)) of participants in DARE.

**Type 1 diabetes genetic risk score generation (T1DGRS)**

All variants were present in the imputed genotype data determined from UK Biobank Affymetrix Axiom Array data (3). This contained 35 HLA region SNPs: 14 tag SNPs for HLA DQA1-DQB1 haplotypes and 21 other HLA region SNPs not representing DR-DQ haplotypes. The remaining 32 SNPs were for non-HLA, each independently associated with T1D. All variants were present in the imputed genotype data in UK Biobank. A calculated score at the DR-DQ locus (4) was added to a score for the remaining variants generated by summing the effective allele dosage of each variant multiplied by the natural log (ln) of the odds ratio.

**C-peptide**

C-peptide was measured on stored non fasting EDTA at DARE recruitment (non-fasting, participants recruited after January 2010) and from July 2014, with specific separate participant consent on subsequent routine non-fasting EDTA samples sent as part of routine care. Consent to measure C-peptide on routine clinical samples was sought from all new study participants from May 2014 and all existing insulin treated participants from July 2014. Laboratory analysis was performed by the Academic department of Blood Sciences Department at the Royal Devon and Exeter Hospital using an electrochemiluminescence immunoassay on a Roche Diagnostics E170 analyser (Roche, Mannheim, Germany). Where more than one C-peptide value was available (70% of participants, with a median three samples each), the median C-peptide value was used. The median diabetes duration at C-peptide assessment was 14 years.

***T1D probability estimate: formula for calculating T1D model score***

a) BMI + Age at diagnosis (AD)

$$= 37.94 + (-5.09 \times \log(AD)) + (-6.34 \times \log(BMI))$$

b) BMI + Age at diagnosis+ Sex + HDL + Total Cholesterol (CHO)+ Triglycerides (TRIG)
Nb all variables standardized against participants with diabetes meeting eligibility criteria = (value-cohort mean)/cohort SD

$$=(-1.4963BMI)+(-1.3358AD)+( 0.2473CHO)+(0.3026Sex)+(0.6999HDL)+(-0.5322TRIG)-4.0927$$

**To convert score to probability**

**T1D probability = (exp(T1D model score))/(1+exp(T1D model score))**

### ESM figures

***ESM table 1: Characteristics of all evaluated participants with diabetes from UK Biobank and DARE split by study and insulin treatment status.*** *Data are mean (SD) or n (%). For UK Biobank in minority of participants where lipid data is missing lipid and clinical features model T1D probability has been replaced by clinical features probability to allow all participants to be classified.*

| | **UK Biobank** | | **DARE** | |
|---|---|---|---|---|
| | **Currently insulin treated n=4,845** | **Currently non-insulin treated n=18,253** | **Currently insulin treated n=784** | **Currently non-insulin treated n=457** |
| **Age at diagnosis (years)** | 41.1 (14.8) | 53.8 (10.2) | 39.9 (18.2) | 57.0 (11.4) |
| **Diabetes duration at recruitment (years)** | 17.4 (12.9) | 6.5 (8.4) | 19.4 (12.0) | 10.3 (6.3) |
| **Current BMI (kg/m$^2$)** | 30.6 (6.1) | 31.6 (5.7) | 29.7 (6.2) | 31.2 (6.0) |
| **Sex (Male)** | 2,986 (62%) | 11,769 (64%) | 459 (59%) | 287 (63%) |
| **Currently treated with oral hyoglycaemic agent** | 2,378 (49%) | 12,012 (66%) | 357 (46%) | 396 (87%) |
| **Insulin treatment within a year of diagnosis** | 2,432 (50%) | 287 (2%) | 390 (50%) | 2 (0%) |
| **Clinical model T1D probability (%)** | 24 (34) | 5 (15) | 31 (39) | 4 (8) |
| **Lipid model T1D probability (%)** | 26 (34) | 6 (14) | Not available | Not available |
| **Type 1 diabetes genetic risk score (T1DGRS)** | 11.58 (2.94) | 10.16 (2.41) | Not available | Not available |
| **C-peptide** | Not available | Not available | 652 (779) | 1610 (884) |
| **T1D codes >0.5 T2D** | 676 (30%) *Total with codes n=2,250* | 12 (0.2%) *Total with codes n=8,241* | Not available | Not available |
| **Self reported type 1 diabetes** | 320 (48%) *Total responses n=667* | 17 (1%) *Total responses n=2,809* | 335 (43%) | 0 (0%) |
| **White European** | 3534 (73%) | 13,085 (72%) | 774 (99%) | 450 (98%) |

**ESM Table 2*****: Comparative performance of approaches classifying all insulin treated white European participants with diabetes in UK Biobank combined with current oral hypoglycaemic agent (OHA)****. Cases are classified as type 1 diabetes if they meet the stated criteria, and are otherwise classified as type 2 diabetes. Results ranked by accuracy (total percentage correctly classified) then type 1 diabetes PPV. Brackets signify 95% CI, Positive predictive value (PPV).*

| | Called Type 1 Diabetes | | | Called Type 2 Diabetes | | | |
|---|---|---|---|---|---|---|---|
| **Approach** | **(n)** | **PPV** | **Sensitivity** | **(n)** | **PPV** | **Sensitivity** | **Accuracy** |
| **Lipid model probability ≥12% and no OHA and insulin within a year of diagnosis** | 1062 | 89% (86-92) | 73% (71-76) | 2472 | 86% (83-88) | 95% (94-96) | 87% |
| **Clinical model probability ≥12% and no OHA and insulin within a year of diagnosis** | 960 | 91% (88-94) | 68% (65-70) | 2574 | 84% (81-86) | 96% (95-97) | 86% |
| **ICD algorithm and no OHA and insulin within a year of diagnosis** | 1021 | 89% (86-92) | 70% (68-73) | 2513 | 85% (82-87) | 95% (94-96) | 86% |
| **Lipid model probability ≥12% and no OHA** | 1245 | 83% (80-86) | 79% (77-82) | 2289 | 88% (86-91) | 90% (89-92) | 86% |
| **UK Biobank probable & possible type 1 diabetes and no OHA and insulin within a year of diagnosis** | 886 | 93% (90-96) | 63% (61-66) | 2648 | 82% (80-85) | 97% (96-98) | 85% |
| **Clinical model probability ≥12% and no OHA** | 1112 | 85% (82-88) | 73% (70-75) | 2422 | 85% (83-88) | 92% (91-93) | 85% |
| **ICD algorithm and no OHA** | 1177 | 82% (79-86) | 75% (73-77) | 2357 | 86% (84-89) | 91% (90-92) | 85% |
| **Age diabetes diagnosed <35 years and no OHA and insulin within a year of diagnosis** | 782 | 96% (93-99) | 58% (55-61) | 2752 | 80% (78-83) | 99% (98-99) | 84% |
| **UK Biobank probable & possible type 1 diabetes and no OHA** | 1006 | 87% (84-90) | 68% (65-70) | 2528 | 83% (81-86) | 94% (93-95) | 84% |
| **Age diabetes diagnosed <35 years and no OHA** | 873 | 90% (87-94) | 61% (58-64) | 2661 | 81% (79-84) | 96% (95-97) | 83% |
| **BMI ≤25 (kg/m2) and no OHA and insulin within a year of diagnosis** | 476 | 82% (77-87) | 30% (28-33) | 3058 | 70% (68-73) | 96% (95-97) | 72% |
| **BMI ≤25 (kg/m2) and no OHA** | 567 | 76% (71-80) | 33% (31-36) | 2967 | 71% (68-73) | 94% (93-95) | 72% |

**ESM Table 3:** ***Comparative performance of approaches classifying all insulin treated white European participants with diabetes in UK Biobank with linked diabetes diagnosis codes.*** *Cases are classified as type 1 diabetes if they meet the stated criteria, and are otherwise classified as type 2 diabetes. Results ranked by accuracy (total percentage correctly classified) then type 1 diabetes PPV. Brackets signify 95% CI,* Positive predictive value (PPV).

| | **Called Type 1 Diabetes** | | | **Called Type 2 Diabetes** | | | |
|---|---|---|---|---|---|---|---|
| **Approach** | **(n)** | **PPV** | **Sensitivity** | **(n)** | **PPV** | **Sensitivity** | **Accuracy** |
| **Lipid model probability ≥12% and insulin within year of diagnosis** | 536 | 86% (82-90) | 80% (77-83) | 1108 | 90% (86-93) | 93% (91-95) | 88% |
| **Clinical model probability ≥12% and insulin treated within a year** | 484 | 88% (83-92) | 74% (70-77) | 1160 | 87% (83-91) | 94% (93-96) | 87% |
| **UK Biobank (probable + possible T1D) and insulin treated within a year** | 445 | 88% (84-93) | 68% (65-72) | 1199 | 85% (81-89) | 95% (94-96) | 86% |
| **Lipid model probability ≥12%** | 680 | 75% (70-79) | 88% (86-91) | 964 | 93% (89-97) | 84% (82-86) | 86% |
| **GP diagnostic codes algorithm and insulin treated within a year** | 529 | 81% (76-85) | 74% (70-78) | 1115 | 87% (83-91) | 90% (89-92) | 85% |
| **Clinical model probability ≥12%** | 609 | 77% (72-81) | 81% (78-84) | 1035 | 89% (85-93) | 87% (85-89) | 85% |
| **Age at diagnosis <35 years and insulin treated within a year** | 392 | 91% (86-96) | 62% (58-66) | 1252 | 82% (79-86) | 97% (96-98) | 84% |
| **GP diagnostic codes alone** | 520 | 79% (75-84) | 72% (68-75) | 1124 | 85% (82-89) | 90% (88-92) | 84% |
| **GP diagnostic codes and age and insulin treated within a year** | 603 | 76% (72-81) | 80% (77-83) | 1041 | 89% (85-93) | 87% (85-89) | 84% |
| **GP diagnostic codes alone and insulin treated within a year** | 477 | 81% (77-86) | 67% (64-71) | 1167 | 84% (80-88) | 92% (90-93) | 83% |
| **UK Biobank (probable + possible T1D)** | 548 | 77% (73-82) | 74% (70-77) | 1096 | 86% (82-90) | 88% (86-90) | 83% |
| **Age at diagnosis <35 years** | 481 | 79% (74-84) | 66% (62-70) | 1163 | 83% (79-87) | 91% (89-92) | 82% |
| **GP diagnostic code algorithm** | 627 | 73% (68-78) | 79% (76-83) | 1017 | 88% (84-92) | 84% (82-86) | 82% |
| **GP diagnostic codes and age** | 760 | 67% (63-71) | 88% (86-91) | 884 | 92% (88-97) | 76% (74-79) | 81% |

***Supplementary Table 4: Comparative performance of approach's classifying type 2 diabetes in UK Biobank and Dare in insulin treated participants****. Cases are classified as T2D if they meet the standard criteria. Results ranked in UK Biobank by T2D PPV then* Sensitivity for identifying type 2 diabetes*. Analysis in UK Biobank restricted to White Europeans. Brackets signify 95% CI, Positive predictive value (PPV).*

| | **UK BIOBANK** | | **DARE** | |
|---|---|---|---|---|
| **Approach** | **PPV of cases called type 2 diabetes** | **Sensitivity for identifying type 2 diabetes** | **PPV of cases called type 2 diabetes** | **Sensitivity for identifying type 2 diabetes** |
| **UK Biobank probable type 2 diabetes** | 100% (96-100) | 17% (15-18) | 100% (97-100) | 26% (18-34) |
| **Non metformin oral hypoglycaemic agent** | 100% (96-100) | 17% (15-18) | 100% (97-100) | 23% (15-31) |
| **UK Biobank probable type 2 diabetes and no insulin within a year of diagnosis** | 100% (97-100) | 13% (12-15) | 100% (95-100) | 17% (8-25) |
| **Clinical model probability <5% and no insulin within year of diagnosis** | 97% (93-100) | 48% (46-49) | 95% (92-98) | 45% (38-52) |
| **Lipid model probability <5% and no insulin within year of diagnosis** | 97% (93-100) | 47% (45-48) | n/a | n/a |
| **Lipid model probability <5%** | 95% (92-98) | 66% (64-68) | n/a | n/a |
| **BMI ≥28 (kg/m2) and insulin within a year of diagnosis** | 95% (92-98) | 56% (54-58) | 93% (90-96) | 49% (43-55) |
| **Clinical model probability <5%** | 94% (91-97) | 67% (65-69) | 91% (88-94) | 64% (59-69) |
| **Age diabetes diagnosed ≥40 years and no insulin within a year of diagnosis** | 93% (90-96) | 58% (56-60) | 91% (87-94) | 56% (51-62) |
| **Age diabetes diagnosed ≥40 years** | 85% (82-88) | 83% (80-85) | 83% (79-86) | 81% (77-85) |
| **BMI ≥28 (kg/m2)** | 79% (77-82) | 78% (76-81) | 74% (70-78) | 70% (65-74) |

**ESM Table 5:** ***Comparative performance of approaches classifying all insulin treated participants with diabetes in UK Biobank****. Cases are classified as type 1 diabetes if they meet the stated criteria, and are otherwise classified as type 2 diabetes. Results ranked by accuracy (total percentage correctly classified) then type 1 diabetes PPV. Brackets signify 95% CI,* Positive predictive value (PPV).

| | Called Type 1 Diabetes | | | Called Type 2 Diabetes | | | |
|---|---|---|---|---|---|---|---|
| **Approach** | **(n)** | **PPV** | **Sensitivity** | **(n)** | **PPV** | **Sensitivity** | **Accuracy** |
| **Lipid model probability ≥12% and insulin within year of diagnosis** | 1480 | 84% (81-86) | 87% (86-89) | 3365 | 95% (92-97) | 93% (92-94) | 91% |
| **UK Biobank probable & possible type 1 diabetes and insulin within a year of diagnosis** | 1208 | 88% (85-91) | 75% (73-77) | 3637 | 90% (88-92) | 96% (95-96) | 90% |
| **Clinical model probability ≥12% and insulin a within year of diagnosis** | 1353 | 85% (82-88) | 81% (79-83) | 3492 | 92% (90-94) | 94% (93-95) | 90% |
| **Age diabetes diagnosed <35 years and insulin within a year of diagnosis** | 1088 | 90% (86-93) | 69% (66-71) | 3757 | 88% (86-90) | 97% (96-97) | 89% |
| **UK Biobank probable & possible type 1 diabetes** | 1517 | 74% (71-77) | 79% (77-82) | 3328 | 91% (89-93) | 89% (88-90) | 86% |
| **Self reported diabetes type (n=667 available)** | 320 | 74% (67-81) | 93% (90-96) | 347 | 95% (88-101) | 80% (76-83) | 85% |
| **Age diabetes diagnosed <35 years** | 1376 | 74% (70-77) | 72% (69-74) | 3469 | 88% (86-91) | 89% (88-90) | 84% |
| **Lipid model probability ≥12%** | 1970 | 67% (64-70) | 93% (92-94) | 2875 | 97% (94-99) | 81% (80-82) | 84% |
| **Clinical model probability ≥12%** | 1791 | 67% (64-70) | 84% (82-86) | 3054 | 93% (90-95) | 83% (81-84) | 83% |
| **BMI ≤25 (kg/m2) and insulin within a year of diagnosis** | 652 | 76% (71-80) | 35% (32-37) | 4193 | 78% (76-80) | 95% (95-96) | 78% |
| **BMI ≤25 (kg/m2)** | 872 | 62% (58-67) | 38% (36-41) | 3973 | 78% (76-80) | 90% (89-91) | 75% |

**ESM Figure 1a**

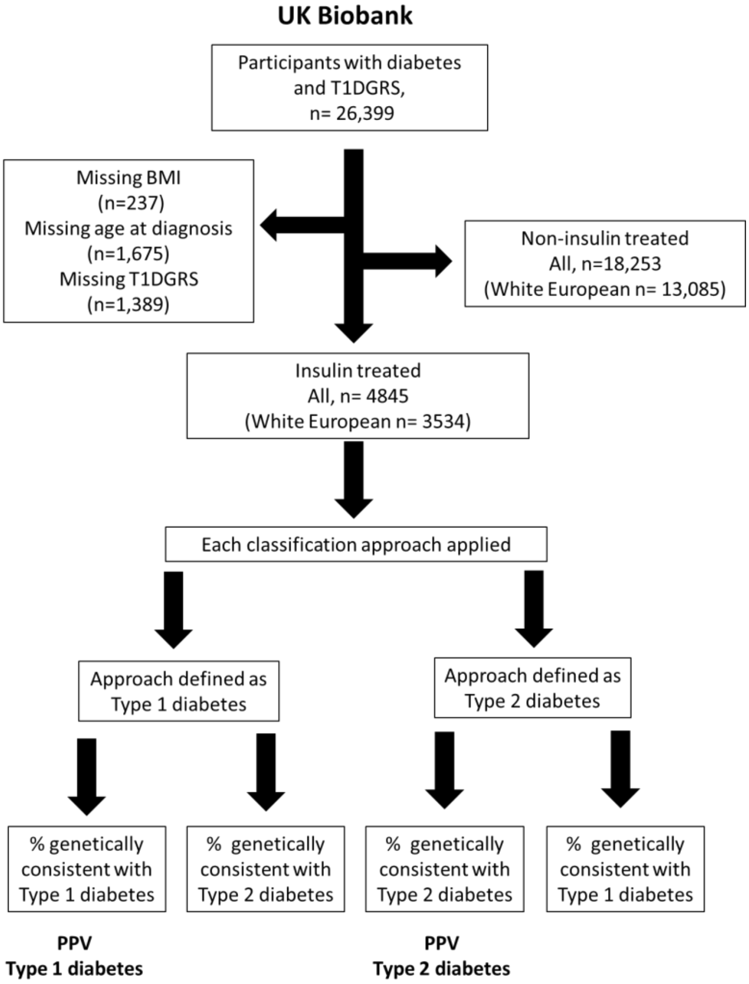


***Study flow diagram for UK Biobank.***

### ESM Figure 1b

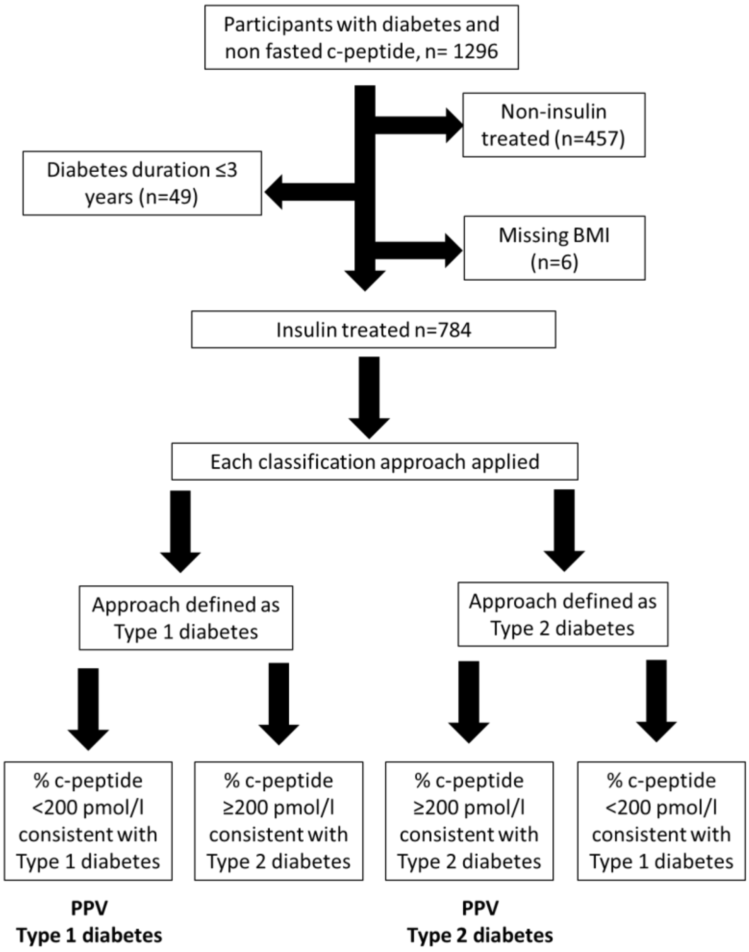


*Study flow diagram for DARE.*

*ESM* **Figure 2.**

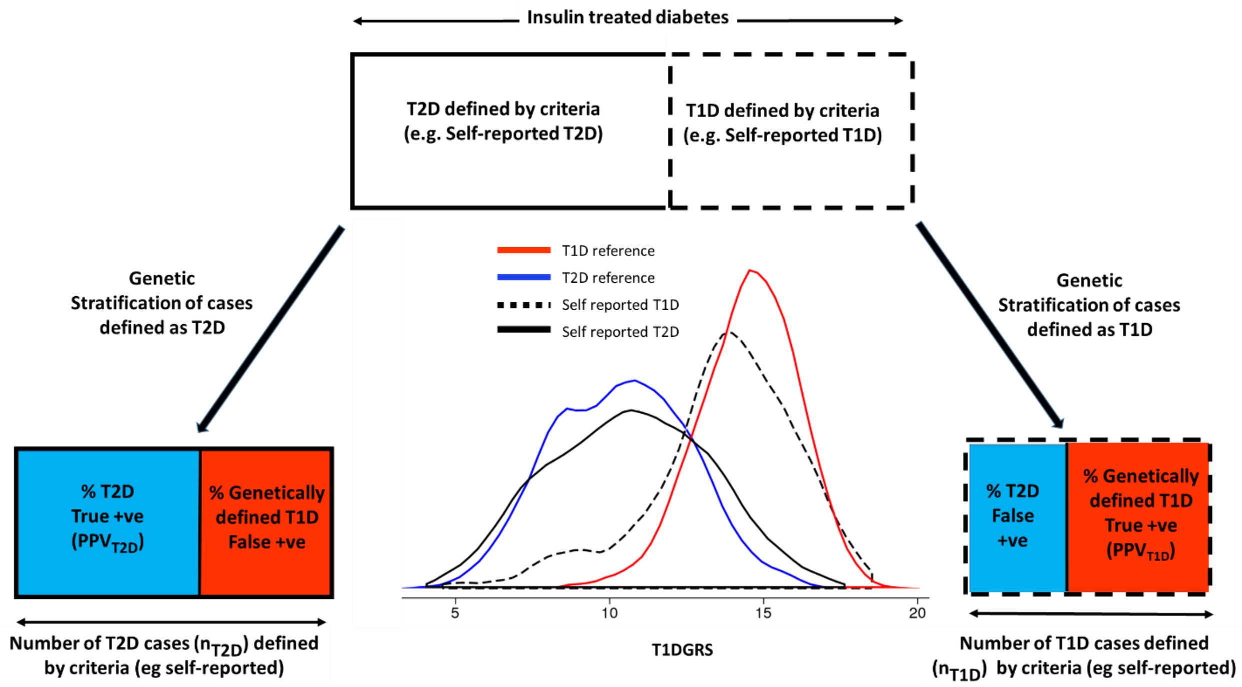


$$Accuracy = \left|\frac{(PPV_{T1D} \times n_{T1D}) + (PPV_{T2D} \times n_{T2D})}{(n_{T1D} + n_{T2D})}\right|.$$

***Overview of genetic determination of type 1 diabetes PPV, type 2 diabetes PPV and accuracy****. A clinical classification approach is used to divide insulin treated diabetes cases into type 1 and type 2 diabetes: self reported diabetes type is shown as an example. The distribution of T1DGRS of cases defined by the clinical approach as either T1D or T2D are separately evaluated against the T1DGRS of reference type 1 and type 2 diabetes cases to genetically estimate the proportion of T1D and T2D within each cohort. The genetically estimated proportion of T1D in those clinically defined as T1D by an approach is the positive predictive value (PPV) for type 1 diabetes. The PPV for type 2 diabetes is calculated as 1-the genetically estimated proportion of T1D in those clinically defined as T2D by an approach.*

### ESM references


1. Welsh C, Celis-Morales CA, Brown R, Mackay DF, Lewsey J, Mark PB, Gray SR, Ferguson LD, Anderson JJ, Lyall DM, Cleland JG, Jhund PS, Gill JMR, Pell JP, Sattar N, Welsh P. Comparison of Conventional Lipoprotein Tests and Apolipoproteins in the Prediction of Cardiovascular Disease. Circulation 2019;140:542-552
2. Elliott P, Peakman TC, Biobank UK. The UK Biobank sample handling and storage protocol for the collection, processing and archiving of human blood and urine. International journal of epidemiology 2008;37:234-244
3. Bycroft C, Freeman C, Petkova D, Band G, Elliott LT, Sharp K, Motyer A, Vukcevic D, Delaneau O, O'Connell J, Cortes A, Welsh S, Young A, Effingham M, McVean G, Leslie S, Allen N, Donnelly P, Marchini J. The UK Biobank resource with deep phenotyping and genomic data. Nature 2018;562:203-209
4. Sharp SA, Rich SS, Wood AR, Jones SE, Beaumont RN, Harrison JW, Schneider DA, Locke JM, Tyrrell J, Weedon MN, Hagopian WA, Oram RA. Development and Standardization of an Improved Type 1 Diabetes Genetic Risk Score for Use in Newborn Screening and Incident Diagnosis. Diabetes care 2019;42:200-207
